# ChatGPT takes the FCPS exam in Internal Medicine

**DOI:** 10.1101/2024.06.11.24308808

**Authors:** Hina Qazi, Syed Ahsan Ali, Muhammad Irfan, M. A. Rehman Siddiqui

## Abstract

Large language models (LLMs) have exhibited remarkable proficiency in clinical knowledge, encompassing diagnostic medicine, and have been tested on questions related to medical licensing examinations. ChatGPT has recently gained popularity because of its ability to generate human-like responses when presented with exam questions. It has been tested on multiple undergraduate and subspecialty exams and the results have been mixed. We aim to test ChatGPT on questions mirroring the standards of the FCPS exam, the highest medical qualification in Pakistan.

We used 111 randomly chosen MCQs of internal medicine of FCPS level in the form of a text prompt, thrice on 3 consecutive days. The average of the three answers was taken as the final response. The responses were recorded and compared to the answers given by subject experts. Agreement between the two was assessed using the Chi-square test and Cohen’s Kappa with 0.75 Kappa as an acceptable agreement. Univariate regression analysis was done for the effect of subspeciality, word count, and case scenarios in the success of ChatGPT.. Post-risk stratification chi-square and kappa statistics were applied.

ChatGPT 4.0 scored 73% (69%-74%). Although close to the passing criteria, it could not clear the FCPS exam. Question characteristics and subspecialties did not affect the ChatGPT responses statistically. ChatGPT shows a high concordance between its responses indicating sound knowledge and a high reliability.

This study’s findings underline the necessity for caution in over-reliance on AI for critical clinical decisions without human oversight. Creating specialized models tailored for medical education could provide a viable solution to this problem.

**Author Summary:** Artificial intelligence is the future of the world. Since the launch of ChatGPT in 2014, it become one of the most widely used application for people in all fields of life. A wave of excitement was felt among the medical community when the chatbot was announced to have cleared the USMLE exams. Here, we have tested ChatGPT on MCQs mirroring the standard of FCPS exam questions. The FCPS is the highest medical qualification in Pakistan. We found that with a vast data base, ChatGPT could not clear the exam in all of the three attempts taken by it. ChatGPT, however, scored a near passing score indicating a relatively sound knowledge.

We found ChatGPT to be a consistent LLM for complex medical scenarios faced by doctors in their daily lives irrespective of the subspecialty, length or word count of the questions. Although ChatGPT did not pass the FCPS exam, its answers displayed a high level of consistency, indicating a solid understanding of internal medicine. This demonstrates the potential of AI to support and improve medical education and healthcare services in near future.

## Introduction

Artificial Intelligence (AI) is increasingly establishing its role in a variety of specialties, including the medical field. It has revolutionized the human approach to various tasks and problems. In recent years, AI has been tried by medical professionals in diagnosis, precision medicine, and research(1). One of the remarkable inventions of AI is ChatGPT (Chat Generative Pre-trained Transformer) which is a natural language processing (NLP) Chatbot driven by AI technology. It is a state-of-the-art language model designed to generate human-like text based on the input it receives. This large language model (LLM) can answer a variety of questions and reply to ‘prompts’ on request. ChatGPT was released in November 2022 and crossed the one million user mark in just five days after it was made public. (2). It has gained a high surge of interest in the medical field in the last few months, with endless speculations about its predictive capabilities, and at least 10 authorships in peer-reviewed scientific journals(3).

AI has been indicated as a safe and effective tool for triage in ER demonstrating high sensitivity in deciding the need for patient admission and providing comprehensive diagnoses, in addition to offering treatment strategies(4). ChatGPT exhibits promising potential in contributing to clinical decision-making skills. A recent study by Rao et al. reported clinical accuracy of up to 71.7% for ChatGPT in making clinical judgments, diagnosis, and management decisions. However, it lagged in listing appropriate differential diagnoses which are the essence of a physician’s role (5). It has been studied as a feasible tool for radiologic decision-making, with the potential to improve clinical workflow and responsible use of radiology services. (6)

Recently, more healthcare consumers have turned to the internet to seek health-related advice. It is of a major concern whether the information accessed is accurate and comparable to that of a physician (7). When comparing the safety of responses of ChatGPT to eye-related patient queries to that of qualified ophthalmologists, the AI did not differ much from humans in the likelihood of causing harm(8).

ChatGPT has rapidly demonstrated its ability to answer exam-style questions and provide explanations, raising questions about its potential role in education and assessment. (9) There have been several recent studies in which the AI chatbot has taken various exams in different specialties. It is seen to be capable of achieving a passing grade when tested on exams such as the United States Medical Licensing Examination and the European Exam in Core Cardiology (10, 11).

ChatGPT was also able to pass BLS and ACLS exams with the questions being presented as open-ended questions with outstanding results. However, it failed when the same questions were presented in a multiple-choice question format(12). These facts taken together make a compelling case for the potential applications of ChatGPT as an interactive medical education tool to support learning. However, there are some studies indicating that it did not perform well in exams like Taiwan’s Family Medicine Board Exam and AHA MCQs(12, 13). When pitted against practice questions of specialized board exams like the neonatal-perinatal medicine board examination, which is taken by practicing pediatricians specializing in neonatology, ChatGPT managed to score only 46%. These suggest further testing of the AI model before incorporating it into medical education and clinical workflows. (14)

The data on the performance of ChaGPT in examinations of low-income countries, with different ethnic and cultural backgrounds, is scarce. This study tested the ability of ChatGPT on exam-style questions that mirror the standards of FCPS theory examinations conducted by CPSP. FCPS examinations are given to candidates enrolled in Fellowship of College of Physicians and Surgeons (FCPS) programs after completion of their training. The theory examination consists of two parts i.e. Paper 1 and Paper 2 with 100 multiple choice questions (MCQs) each covering the thorough knowledge of internal medicine, testing pathophysiology, clinical reasoning, and guideline-recommended medical management. The passing criteria is 75%. This study is designed to assess the clinical usefulness of ChatGPT(4.0) and evaluate its performance on questions that lie within the scope of the final exit exam of Fellowship Of College of Physicians and Surgeons (FCPS part 2) examinations conducted by the College of Physicians and Surgeons (CPSP).

## Methods and materials

### ChatGPT-4

This comprehensive study was designed to test ChatGPT (OpenAI; San Francisco, CA)’s performance on questions of FCPS part 2 level in the specialty of internal medicine. We utilized the latest version of ChatGPT, which is based on the GPT-4 architecture, and is an open-access LLM developed by OpenAI (https://openai.com), launched in 2023. As an LLM, it is designed to generate human-like responses. This AI chatbot has been trained on extensive data, including medical texts and journals, up to September 2021.

### FCPS 2 exam questions

The FCPS part 2 theory exam consists of multiple-choice questions with 5 choices for each question. As a source of MCQs, publically available exam questions of FCPS part 2 level were used. A total of 111 single-answer MCQs which reflected the standards of the FCPS 2 exam were obtained and screened. Approximately ten questions from each subspecialty namely endocrinology, cardiology, nephrology, neurology, pulmonology, infectious diseases, hematology, and oncology were selected. Success was defined as ChatGPT achieving the passing criteria of 75% or higher.

### Input to ChatGPT

The selected questions were manually pasted into ChatGPT, preceded by the statement, “Please choose the best answer and explain your reasoning:” in a new separate line. The multiple-choice answers were provided, with each option pasted in a separate new line. A new chat session was started each time to avoid retention bias. About the output from ChatGPT, responses that were incorrect or inconclusive were both considered incorrect for this study. For questions for which ChatGPT provided 2 answers, another prompt was given to choose one best answer, and that answer was taken as the final answer. To evaluate ChatGPT’s performance, each question from the question bank was submitted in the form of a text prompt. To evaluate the consistency and stability of these responses, all questions were submitted to ChatGPT three times on different dates from 15 November to 20 November 2023. The average of those three responses was taken as the final answer. All questions containing visual elements such as clinical images, charts, and tables were excluded. Upon receiving the responses generated by ChatGPT, they were scored against the correct answers provided by the subject experts. Responses from ChatGPT were compared with the answers provided by the subject experts along with their clinical justification. Two subject experts answered these questions in a masked manner. For the questions in which there were differences in opinion, the two subject experts discussed the responses to reach a consensus. Answers on which the subject experts did not have a consensus, were planned to be excluded.

### Statistical Analysis

The statistical analysis was done using SPSS version 23. ChatGPT responses of each attempt and the final key were compared to assess agreement between the two variables using the Chi-square test and Cohen’s Kappa. The Cohen’s kappa value of 0.75 was considered an acceptable agreement. Univariate regression analysis was done for the effect of subspeciality, word count, and case scenarios in the success of ChatGPT. Stratified analysis was done for subspecialties and for answers on which physicians initially agreed. Post-risk stratification chi-square and kappa statistics were applied. A p-value less than 0.05 was considered statistically significant at the confidence interval of 95%.

### Ethical considerations

As this study did not involve any human or animal interaction, no approval from the institutional review board or informed consent was required.

Flowchart:

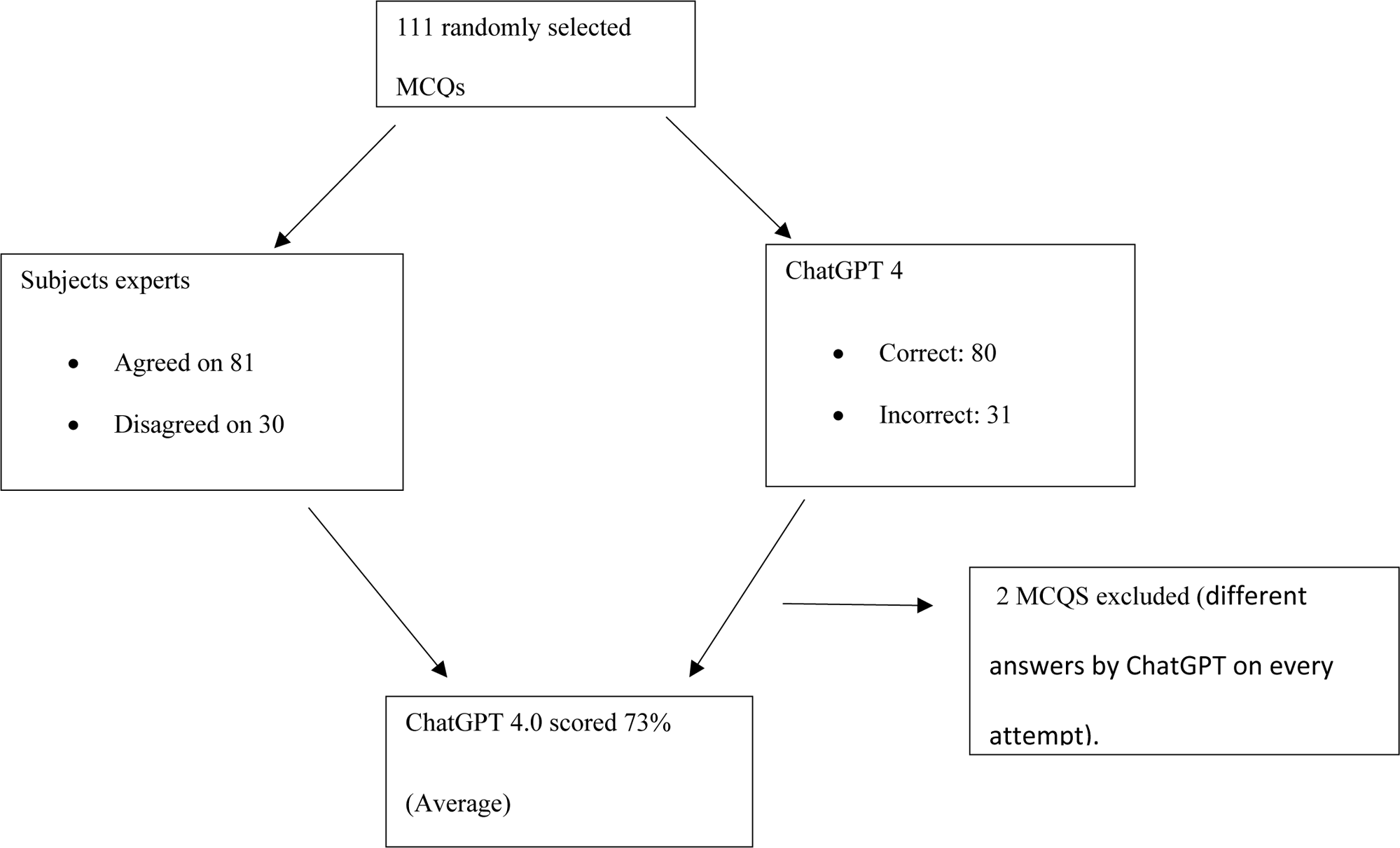

## Results

This study tested the performance of ChatGPT 4.0 on questions that fall within the scope of the FCPS 2 theory examination in internal medicine. Hundred and eleven publically available questions, comprising various subspecialties were tested on ChatGPT (Table 1). Case scenarios comprised a major portion of the exam i.e. 83 questions, while the rest were clinical facts. None of the questions were related to the local policies. The word count of the questions ranged from 8 words to 241 words, with a median of 72 words. With the median as a reference, questions with a word count less than 72 were labeled as short questions and those with more than 72 words were labeled as long questions. Out of 111 questions, subject experts gave similar answers to 81 questions while they differed on 30 questions, indicating the level of difficulty of these questions. They later reached a consensus for every answer after discussion and no disagreement was reported. Their responses were taken as the final key. Two questions were excluded from the statistical analysis as ChatGPT came up with a different answer every time and their average could not be calculated. ChatGPT took three attempts on all questions. In the first attempt. ChatGPT scored 74% (Kappa 0.674).In all three attempts, it could not cross the 75% passing criteria, Of the 81 questions on which the subject experts initially agreed, ChatGPT scored 74%. On the questions, they agreed to after discussion, ChatGPT scored 73%. ChatGPT could not clear its second attempt too, and scored 72.0 %(Kappa 0.672). ChatGPT failed the third attempt with a score of 69.7% (Kappa 0.630). The average of the three attempts was taken as the final score and was 73% (Kappa 0.675)(p 0.001).

**Table 1:** Qualitative analysis of question characteristics.

| **Subspecialties** | **Frequency** | **Percent** |
| --- | --- | --- |
| Nephrology | 9 | 8.3 |
| Neurology | 12 | 11.1 |
| Cardiology | 10 | 9.3 |
| Dermatology | 1 | 0.9 |
| Endocrinology | 13 | 12 |
| Forensic | 1 | 0.9 |
| Gastroenterology | 14 | 13 |
| Hematology/oncology | 15 | 13.9 |
| Infectious Diseases | 9 | 8.3 |
| Pulmonology | 11 | 10.2 |
| Rheumatology | 13 | 12 |
| **Questions on physicians Initially Agreed** |  |  |
| No | 29 | 26.9 |
| Yes | 79 | 73.1 |
| **Question length** |  |  |
| Short | 53 | 49.1 |
| Long | 55 | 50.9 |
| **Case scenarios** |  |  |
| No | 25 | 23.1 |
| Yes | 83 | 76.9 |
| **Basic medical facts** |  |  |
| No | 85 | 78.7 |
| Yes | 23 | 21.3 |
| Total | 108 | 100.0 |

Statistically, the consistency between attempts was seen by assessing the inter-attempt Kappa agreement and the chi-square test. The first attempt when compared to the second attempt showed an almost perfect agreement of 0.99 or 99%. The second attempt when compared to the third attempt gave an agreement of 0.92 or 92%.The third attempt when compared to the first attempt showed concordance of 88% or an agreement of 0.88.

On univariate regression analysis, no statistical difference was found concerning subspecialty, word count, and case scenarios in the success of ChatGPT (p 0.175). Therefore, binary logistic regression analysis could not be done. Stratified analysis was done for subspecialties and for answers on which physicians initially agreed. Post-risk stratification chi-square and kappa statistics were applied. Pulmonology had the highest agreement of 100% while nephrology had the lowest of 8.3% but this difference was statistically non-significant. The qualitative analysis of the responses by ChatGPT is given in Table 2.

**Table 2:** Qualitative analysis of responses generated by ChatGPT.

|  |  |  | Final Key by subject experts |  |  |  |  | Total |
| --- | --- | --- | --- | --- | --- | --- | --- | --- |
|  |  |  | Option<br>A | Option B | Option<br>C | Option<br>D | Option<br>E |  |
| **Answers by**<br>**ChatGPT**<br>**(Average of 3**<br>**attempts)** | **Option A** | Percentages<br>within an<br>average<br>attempt | 84.2% | 10.5% | 5.3% | 0.0% | 0.0% | 100.0% |
|  | **Option B** | Percentages<br>within an<br>average<br>attempt | 10.3% | 65.5% | 3.4% | 6.9% | 13.8% | 100.0% |
|  | **Option C** | Percentages<br>within an<br>average<br>attempt | 9.1% | 0.0% | 86.4% | 0.0% | 4.5% | 100.0% |
|  | **Option D** | Percentages<br>within an<br>average<br>attempt | 22.2% | 0.0% | 0.0% | 66.7% | 11.1% | 100.0% |
|  | **Option E** | Percentages<br>within an<br>average<br>attempt | 10.0% | 10.0% | 10.0% | 0.0% | 70.0% | 100.0% |
| **Total percentages** |  |  | 25.0% | 21.3% | 21.3% | 13.0% | 19.4% | 100.0% |

## Discussion

ChatGPT garnered significant media attention and generated a buzz of excitement when it was reported to have cleared the USMLE exam, which is a licensing exam taken by medical undergraduates(10). ChatGPT was also reported to have passed multiple international licensing exams and sub-specialty exams (table 3 and table 4)(Fig.1). Along with enthusiasm, it has sparked skepticism about its accuracy, reliability, and its potential in the field of medicine.

**Fig 1:**
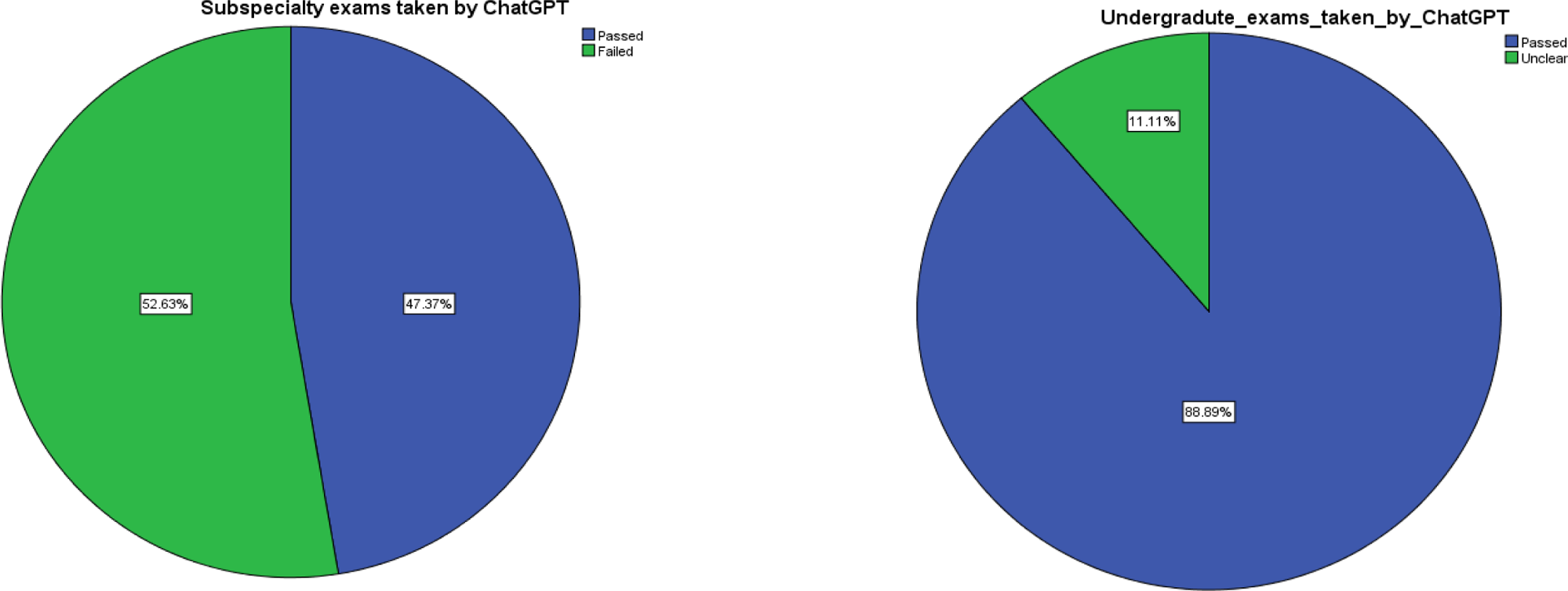
Pie chart indicating the percentages of passed and failed exams taken by ChatGPT

**Table 3:**
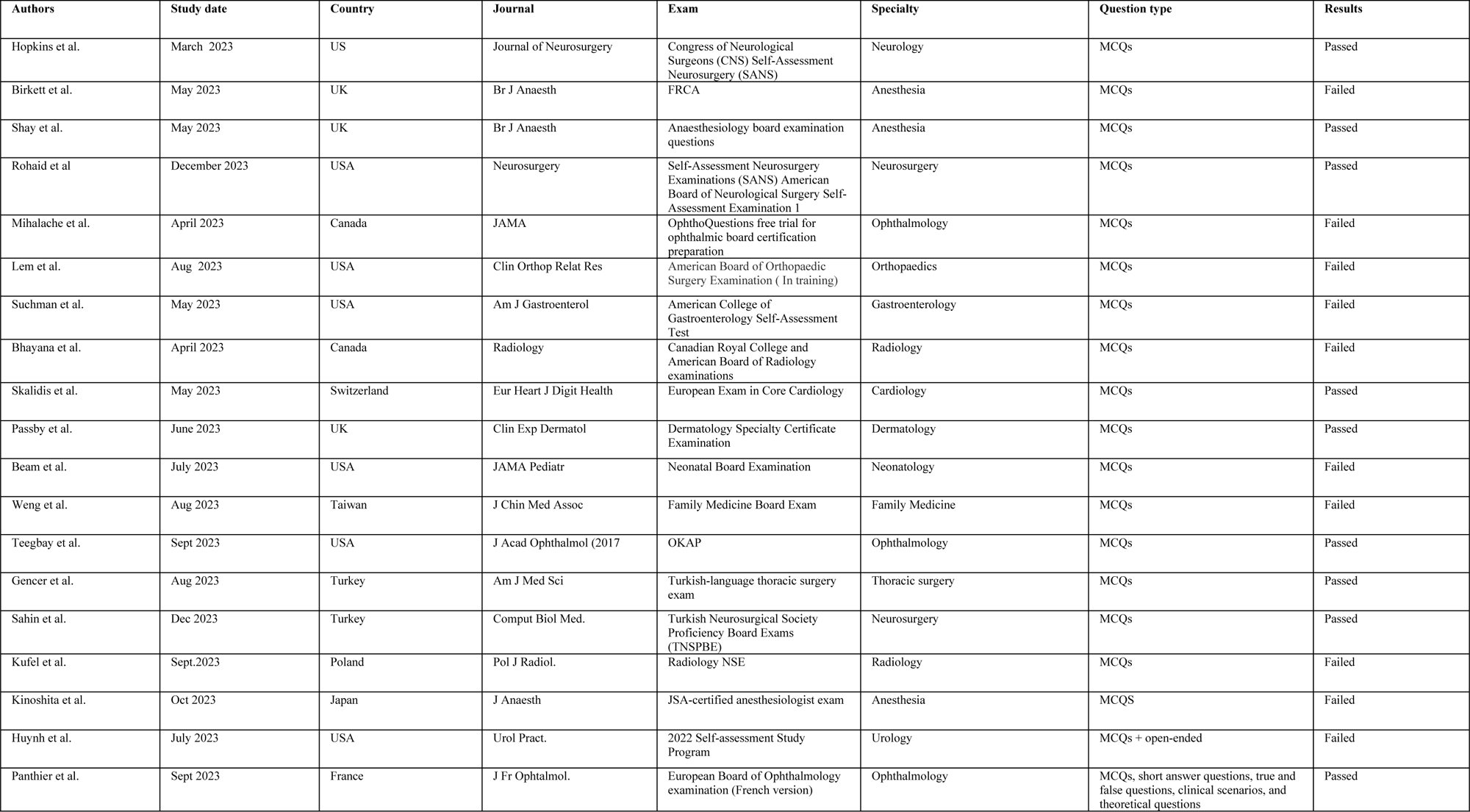
List of sub-specialty exams taken by ChatGPT.

**Table 4:** List of Undergraduate exams taken by ChatGPT.

| Authors | Study date | Country | Journal | Exam | Question type | Results |
| --- | --- | --- | --- | --- | --- | --- |
| Gilson et al. | Feb 2023 | USA | JMIR Med Educ | USMLE | MCQs | Pass |
| Kung et al. | Feb 2023 | USA | PLOS Digit Health | USMLE | MCQs | Pass |
| Aljindan et al. | Sept 2023 | Saudia | Cureus | Saudi Medical Licensing Exam | MCQs | Pass |
| Wang et al. | Aug 2023 | China | J Med Syst | Chinese National Medical Licensing Examination | MCQs | Below performing students |
| Huang et al. | Oct 2023 | Taiwan | Healthcare (Basel) | Registered Nurse License Exam | MCQs | Pass |
| Rosol et al. | Nov 2023 | Poland | Sci Rep. | Polish Medical Final Examination | MCQs | Pass |
| Kasai et al. | April 2023 | Japan | Springer | Japanese Medical Licensing Examinations | MCQs | Pass |
| Lai et al. | Sept 2023 | UK | Front Med (Lausanne). | UKMLA | MCQs | Pass |
| Bonetti et al. | July 2023 | Italy | Ann Biomed Eng. | Italian Residency Admission | MCQs | Pass |

The results of this study indicate that while ChatGPT demonstrates a reasonable level of knowledge in internal medicine, as evidenced by its nearly passing scores in the FCPS 2 examinations, it does not yet meet the benchmark for clinical reliability. The FCPS exam is a highly specialized exam taken by postgraduate trainees after the completion of their 4-year training, to be a qualified consultant in their field. Other studies have reported similar results, where ChatGPT failed to clear specialty exams (Table 2). For example, ChatGPT failed the Taiwan Family Medicine exam (13). There could be two reasons as to why it failed. Either it could not accurately answer family medicine-related questions or it was unable to respond correctly to prompts in Chinese language. ChatGPT could also not perform well on the American Academy of Gastroenterology exam questions scoring 62.4 percent compared to the required 70 percent to pass the exams(16). Similarly, ChatGPT showed poorer responses on American Board of Orthopedic Surgery style questions when compared with responses from in-training residents, scoring only 47% (17).

Assessing the performance of ChatGPT on FCPS questions, it consistently failed all three attempts, with the lowest score on the last attempt. Eighty-three MCQs in this study were based on clinical scenarios and ChatGPT answered 62 of them correctly, indicating a relatively sound knowledge of diagnosing medical conditions, interpreting diagnostic testing and lab reports providing appropriate management plans. Question characteristics such as word count, case scenarios, and clinical facts statistically did not seem to affect the performance of ChatGPT. This finding is in line with Rohaid Ali et al who tested ChatGPT-4 on neurosurgery questions and reported no effect of responses on the word count of the questions(18).

There could be multiple explanations as to why ChatGPT failed the FCPS exam. ChatGPT was never trained specifically on medical literature and was developed as a general interactive LLM. The AI model’s training on historical data up to September 2021 may not encompass the most recent medical guidelines and research, and its inability to access paid content of medical journals could have led to less optimal responses. ChatGPT gathers data from various sources, some of which may be nonmedical, semi-medical, or outdated, resulting in inaccurate responses. The way it works is to predict the next most suitable word in a string, generating a likely reply based on existing data, without any regard to factual accuracy. The model lacks inherent comprehension of any subject or matter. ChatGPT may have struggled with the MCQ-style format of the FCPS exam. As a chatbot, it may be more suited to answering open-ended questions rather than being given a set of options to choose from. A similar finding was also speculated by Zhu et al.(12). In their study, ChatGPT could clear the open-ended American Heart Association questions but failed on the MCQs of the same. FCPS exam includes a higher proportion of questions that require more nuanced clinical judgment or interpretation, particularly in areas where there might be multiple acceptable approaches, amongst which the trainee has to choose the best. ChatGPT might have found it more challenging.

The impact of ChatGPT in the educational sector has received mixed reactions. While some appreciate the AI’s ability to provide useful insights and demonstrate reasoning skills to students, others point out issues such as the production of inaccurate information, the challenge in interpreting responses, the possibility of bias, and ethical concerns(19).

There are certain limitations of our study. This study only assessed ChatGPT’s performance on MCQs as the theory exam of FCPS only consists of MCQs. This format might not fully capture the AI’s capabilities in other aspects of clinical reasoning, such as case analysis, patient interaction, and practical skills. Secondly, the determination of correct answers was based on the consensus of subject experts. While this method is standard, it introduces a subjective element and could potentially overlook the diversity of acceptable clinical opinions or practices.

The study suggests that future iterations of AI models like ChatGPT 4.0 could benefit from more targeted training, specifically in areas where the model currently underperforms. As we write this, AI can complement, but not replace human expertise. It can also be said that shortly, LLMs when used by internists will make them better internists. On the contrary, some people believe that in the next 10 years, AI chatbots will be primarily making disease diagnoses and doctors will only be reached out for a second opinion. However, this study’s findings underline the necessity for caution in over-reliance on AI for critical clinical decisions without human oversight. Regular updates with recent medical literature and guidelines are crucial to ensure the model’s responses remain current and clinically relevant.

## Conclusion

This cross-sectional study tested randomly selected 111 FCPS-level MCQs on ChatGPT 4.0 on three consecutive attempts. Question characteristics such as word count, case scenarios, and clinical facts did not seem to affect the responses. Although ChatGPT was not able to pass the FCPS exam, it showed a high concordance within its answers indicating relatively sound knowledge of internal medicine and reflecting the potential of AI in assisting and enhancing medical education and healthcare services. We advise caution for those using ChatGPT as a medical education tool. As the advancements in AI technology continue, particularly in areas of clinical interpretation and specific-domain applications of knowledge, it will be interesting to see how this technology continues to improve and how it might best be applied in medical education. Creating specialized models tailored for medical education could provide a viable solution to this problem.

## Data Availability

Data will be available from the principal investigator on reasonable request.

## Acknowledgment

The authors have no financial disclosures or support in this work.

We would like to thank Ms. Khadijah Abid, senior instructor of research at the Department of Ophthalmology and Visual Sciences, AKUH, for helping with the statistical analysis of this study.

## Notes

### Competing Interest Statement

The authors have declared no competing interest.

### Funding Statement

The author(s) received no specific funding for this work.

### Author Declarations

This study did not involve human interaction, so IRB approval was not required.

